# Potential Health Benefits of Eliminating Traffic Emissions in Urban Areas

**DOI:** 10.1101/2021.07.27.21261168

**Authors:** Shahram Heydari, Masoud Asgharian, Frank J Kelly, Rahul Goel

## Abstract

Traffic is one of the major contributors to PM_2.5_ in cities worldwide. Quantifying the role of traffic is an important step towards understanding the impact of transport policies on the possibilities to achieve cleaner air and accompanying health benefits. We carried out a meta-analysis using the World Health Organisation (WHO) database of source apportionment studies of PM_2.5_ concentrations. Specifically, we used a Bayesian meta-regression approach, modelling both overall and traffic-related (tailpipe and non-tailpipe) concentrations simultaneously. We obtained the distributions of expected PM_2.5_ concentrations (posterior densities) of different types for 117 cities worldwide. For each city, we calculated the probabilities of exceeding the WHO-recommended concentration of PM_2.5_ if all of traffic emissions were removed. Using the non-linear Integrated Exposure Response (IER) function of PM_2.5_, we estimated percent reduction in different disease endpoints for a scenario with complete removal of traffic emissions. We found that this results in achieving the WHO-recommended concentration of PM_2.5_ only for a handful of cities that already have low concentrations of pollution. The percentage reduction in prevented mortality for cardiovascular and respiratory diseases increases up to a point (30-40 ug/m^3^), and above this concentration, it flattens off. For Diabetes-related mortality, the percentage reduction in mortality decreases with increasing concentrations—a trend that is opposite to other outcomes. For cities with high concentrations of pollution, the results highlight the need for multi-sectoral strategies to reduce pollution. The IER functions of PM_2.5_ result in diminishing returns of health benefits at high concentrations, and in case of Diabetes, there are even negative returns. The results show the significant effect of the shape of IER functions on health benefits. Overall, despite the diminishing results, a significant burden of deaths can be prevented by policies that aim to reduce traffic emissions even at high concentrations of pollution.

## 1. Introduction

Motorised traffic is growing rapidly in many low- and middle-income countries (LMICs) resulting from increasing ownership of vehicles and rapid urbanisation. In these settings, emissions are still increasing compared to Europe and the United States where emissions have stabilised or are decreasing (Crippa et al., 2018). As expected, fine particulate matter (PM_2.5_) concentrations in East Asia, South Asia, and parts of Sub-Saharan Africa have increased markedly over the past decades and are currently the highest in the world (Apte et al., 2018; Burnett & Cohen, 2020). These concentrations of pollution result in disproportionate share of premature mortality due to cardiovascular and respiratory diseases in LMICs (Burnett & Cohen, 2020). New evidence on the impact of PM_2.5_ pollution on infant mortality (Heft-Neal et al., 2018) and Diabetes (Bowe et al., 2018), now included in the Global Burden of Disease estimates (Stanaway et al., 2018), has added to the previously known health burden of PM_2.5_. Air pollution is one of the key pathways through which transport impacts public health in the cities (Khreis et al., 2017). Health impact studies that focus on reducing the use of motorised travel through a shift to active modes of travel have highlighted the health benefits from reduction in traffic emissions (Mueller et al., 2018; Tainio, 2015).

Several previous studies investigated different aspects of traffic-related air pollution and its health impacts (Chambliss et al., 2014; Liang et al., 2020; Pan et al., 2019; Ramacher et al., 2020; Rodrigues Teixeira et al., 2020; Tong et al., 2020). For example, Pan et al. (2019) estimated potential impacts of electric vehicles on air quality and health endpoints in Houston (USA) in 2040. Tong et al. (2020) examined health effects of PM_2.5_ emissions from on-road vehicles during weekdays and weekends in Beijing, China. Teixeira et al. (2020) estimated the impact of PM emissions from heavy-duty trucks on human health. A review by Health Effects Institute (HEI, 2010) found suggestive evidence of a causal relationship between exposure to traffic-related air pollution and onset of childhood asthma, non-asthma respiratory symptoms, impaired lung function, total and cardiovascular mortality, and cardiovascular morbidity. These studies have all pointed towards the potential of achieving health benefits from reduction in traffic-related air pollution in urban settings.

The potential of gaining health benefits in a city through reduction in traffic emissions depends on the proportion of PM_2.5_ concentrations that is contributed by this sector as well as the total PM_2.5_ burden. Since the dose-response functions are non-linear, with a curve that is steep at low concentrations and flattens towards higher concentrations, there are diminishing returns of reduction in pollution levels at higher concentrations (Apte et al., 2018). The two factors (proportion of traffic and overall PM_2.5_ concentrations) vary greatly across the world (Heydari et al., 2020; Karagulian et al., 2015; WHO, 2018). For example, Heydari et al. (2020) showed that traffic contribution estimates as well as uncertainties around these estimates vary largely across various cities and regions worldwide. Many LMIC cities have high concentrations of pollution because of contributions from multiple sectors, of which transport is only one of them (Guttikunda et al., 2019; WHO, 2018). Many of the high-income countries have achieved cleaner air due to the reduction of emissions across multiple sectors. Based on the WHO source apportionment database (WHO, 2015), employing a population-weighted approach, Karagulian et al. (2015) conducted a systematic review of local source contributions of PM in cities across the world. Using the same database and based on a Bayesian meta-regression approach, Heydari et al. (2020) estimated the expected percentage contribution of traffic to PM_2.5_ and PM_10_, and their respective uncertainties, in various cities and regions worldwide.

The primary goal of this research is to estimate the health benefits that can be gained by reduction in traffic emissions. This is achieved by carrying out a rigorous meta-analysis exercise, with the aim of pooling strength over several previous studies on the concentrations of PM_2.5_ in multiple cities. To this end, in this work we introduce and discuss an analytical framework that can draw valuable inferences regarding the overall (due to all sources) and traffic-related (exhaust and non-exhaust emissions) concentrations of air pollutant concentrations in various locations worldwide from a collection of previous studies. Our specific objectives are summarised as follows:

1. Develop a meta-regression model that simultaneously analyses overall and traffic-related PM_2.5_ concentrations in urban areas based on the previous studies collected in the WHO source apportionment database. Doing so, (i) we can explain variability in the reported concentrations by previous studies; (ii) estimate the magnitude of dependence between overall and traffic-related PM_2.5_ concentrations; and (iii) estimate expected concentrations of PM_2.5_ of different types (traffic-related, non-traffic-related, and overall due to all sources) with their associated uncertainties in multiple cities worldwide.
2. Use the above estimates (specifically, the estimates obtained in step iii) to investigate the potential of achieving cleaner air and preventing premature mortality from multiple disease outcomes through reductions in traffic-related PM_2.5_.

## 2. Materials and methods

### 2.1. Review framework and study selection

We present a meta-analysis of traffic-related PM_2.5_ and overall (due to all sources) PM_2.5_ concentrations reported in the latest World Health Organization (WHO) database on source apportionment studies (WHO, 2015). The WHO database reports overall PM_2.5_ concentrations and the contributions of different source categories (i.e., traffic, industry, domestic fuel burning, natural sources, and unspecified sources of human origins) to particulate matter for various locations. Given the aim of our research, we excluded studies that did not report the share of traffic. Also, we only included studies that were reported from urban areas, excluding other site typologies such as industrial, rural, etc.

Our final dataset includes 182 observations for each of the above-mentioned PM_2.5_ concentration types. These observations are measurements reported by 118 studies (Table A of the supplementary material), corresponding to 117 cities worldwide, from 1987 to 2014. For each study, we obtained traffic-related PM_2.5_ concentrations by multiplying overall PM_2.5_ concentrations by the reported percentage traffic contributions to PM_2.5_. To carry out our quantitative synthesis of previous research, we considered a series of explanatory variables available in the WHO database. These included publication year, study location (city, country, region, and continent), population, geographic coordinates, and estimation method. Another potentially relevant information was whether a study reported sea salt contribution to PM_2.5_. To better capture variability in the data, cities were assigned to 12 different regions mostly according to geographic proximity, and studies conducted by Karagulian et al. (2015) and Heydari et al. (2020). These are North America, Central Europe, East Asia, East/West Africa, Middle East, North-western Europe, Oceania/Japan, South/Central America, South-eastern Asia, Southern Asia, Southwestern Europe, and Western Europe. List of countries in each region are reported in Table B of the supplementary material.

### 2.2. Characteristics of the final data

A summary of the final sample used in our study is reported in Tables 1 and 2. Around 36% of the reported measurements were estimated based on studies conducted in North America or Oceania. Around 10% were from North Western or Western Europe while 23% of the observations were from studies conducted in the rest of Europe. Two-thirds (74.2%) of the measurements were reported after year 2005, and 50% of the observations in the data reported percentage contribution of sea salt to PM_2.5_. In our final data, the reported overall PM_2.5_ concentrations varied largely across cities: from around 12 ug/m^3^ to 97 ug/m^3^, with a mean (and standard deviation) of 35.11 ug/m^3^ (36.97 ug/m^3^) at a global level. Similarly, traffic-related PM_2.5_ varied from 1.10 ug/m^3^ to 64.02 ug/m^3^, with a mean (and standard deviation) of 9.13 ug/m^3^ (12.41 ug/m^3^).

**Table 1.**
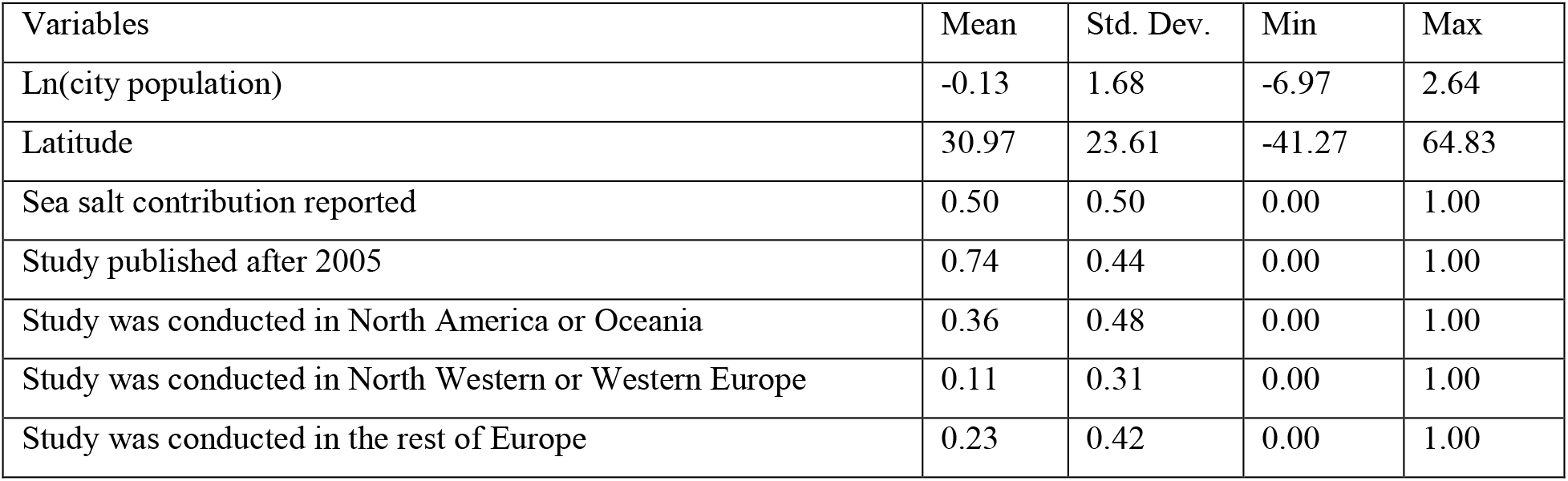
Descriptive statistics of the data

**Table 2.** Distribution of observations in each region

| Regions | Frequency | Percent |
| --- | --- | --- |
| Africa | 1 | 0.55 |
| Central and Eastern Europe | 3 | 1.65 |
| East Asia | 18 | 9.89 |
| Middle East | 5 | 2.75 |
| North America | 57 | 31.32 |
| Northwestern Europe | 12 | 6.59 |
| Oceania/Japan | 9 | 4.95 |
| South/Central America | 12 | 6.59 |
| Southeastern Asia | 9 | 4.95 |
| Southern Asia | 9 | 4.95 |
| Southwestern Europe | 39 | 21.43 |
| Western Europe | 8 | 4.40 |

### 2.3. Meta-regression

We adopted a joint meta-regression approach to identify factors that can explain variations in reported overall PM_2.5_ and traffic-related PM_2.5_ concentrations by previous studies collected in the WHO database. This allowed us to model both outcomes simultaneously through a system-equation approach rather than modelling each outcome separately, improving the reliability of our statistical inferences. We assumed the log-transformed concentrations follow a multivariate normal density. Let ***y***_*ki*_ denote the vector of log-transformed concentrations of *k* different types reported by previous studies *i* (*i* = 1, 2, …, *N*). Here *k* = 2; therefore, ***y*** = (*y*_*1i*_, *y*_*2i*_), where y_1i_, y_2i_ denote PM_2.5_ due to traffic and overall PM_2.5_, respectively. Let ***X***_*k*_ = (*X*_*k1*_, *X*_*k2*_, …, *X*_*km*_) be the vector of *m* explanatory variables (e.g., population) associated with the outcomes of interest (y_1i_, y_2i_) with their respective regression coefficients ***γ* =** (*γ*_*k1*_, *γ*_*k2*_,…, *γ*_*km*_). Let ***η*** = (*η*_*1*_, *η*_*2*_) denote the vector of intercepts corresponding to *y*_*1i*_ and *y*_*2i*_, respectively. Let *R* and *K* denote the scale matrix and the degrees of freedom, respectively, in a Wishart distribution. We can then write

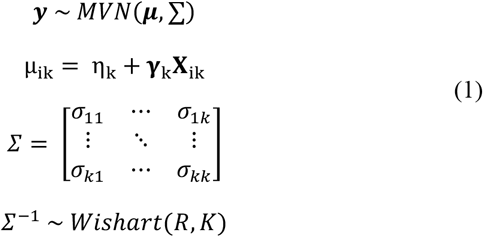

As it can be seen in the above model, the dependency across outcomes is captured through the covariance matrix *Σ*. This specification allowed us to investigate the magnitude of correlation between overall PM_2.5_ and traffic-related PM_2.5_ concentrations.

### 2.4. Prior specification and model computation

Normally distributed non-informative priors, normal (0,100), were placed on the regression coefficients. As it is common in multivariate settings, we specified a Wishart distribution for the inverse of covariance matrix *Σ*^*-1*^(Tunaru, 2002) a common approach in joint modelling of correlated outcomes, with *K*=2 (for two correlated outcomes) and a 2×2 scale matrix *R* (*R*[1,1]=*R*[2,2]=0.01 and *R*[1,2]=*R*[2,1]=0), which leads to a non-informative prior specification. For model computation, we employed WinBUGS (Lunn et al., 2000) to draw posterior densities for our Markov chain Monte Carlo simulations running two chains each containing 15,000 iterations. The posterior densities are based on 20,000 samples as the first 5,000 iterations were discarded for convergence requirements. Based on the Gelman-Rubin statistic (Gelman & Rubin, 1992), history plots, and Monte Carlo errors, this number of iterations was sufficient.

### 2.5. Computing probabilities of exceeding the WHO-recommended concentration of PM_2.5_

Our approach allows us to readily estimate the probability of non-traffic-related PM2.5 concentrations exceeding the WHO-recommended concentration of 10 ug/m^3^ (WHO, 2018). The idea here is to obtain probabilities of exceeding 10 ug/m^3^, for various cities available in our data, if traffic emission was eliminated completely. For computing these probabilities, we created an *Mx1* matrix of indicator variables [*I*_*c*_] for each city *c*, where *M* is the total number of cities under investigation. At each iteration of our MCMC simulations and for each city, we compared the expected non-traffic-related PM_2.5_ concentration with the WHO-recommended value of 10 ug/m^3^ as shown in (2). Finally, we averaged the indicator variable value over all iterations to obtain the probabilities of exceeding for each city.

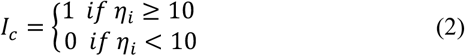

### 2.6. Estimating health benefits of reducing traffic emissions

We estimated the potential of a city to attain the WHO-recommended concentration of PM_2.5_ concentrations (10 ug/m^3^) (WHO, 2018) if all traffic emissions were removed. For this, we estimated the posterior densities of traffic-excluded PM_2.5_ concentrations for each city based on the meta-regression approach discussed in Section 2.3. We then estimated the probability that each posterior mean exceeds 10 ug/m^3^. The higher this probability, the lower the ability of a city to reach the WHO-recommended concentration of PM_2.5_ even from complete removal of traffic emissions. Next, we estimated the percentage reduction in health burden resulting from a complete removal of traffic emissions. To estimate changes in health burden we used Integrated Exposure Response (IER) functions for six disease endpoints that were used in GBD 2017 (Stanaway et al., 2018). These are Ischemic Heart Disease (IHD), Stroke, Chronic Obstructive Pulmonary Disease (COPD), Lung Cancer, Lower Respiratory Infections (LRI), and Type-II Diabetes (Diabetes). For IHD and Stroke, IER is age-specific and, for these, we present calculations for 55-60 years age group for illustration of the method. The relative risk using IER function is calculated as:

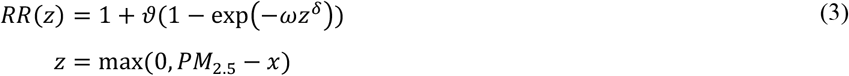

where *ϑ, ω, δ* are parameters specific to each disease end-point and *x* is a counterfactual value below which the assumption is that there are no increased mortality. The value of *x* is obtained from a uniform distribution, representing its uncertainty, with lower and upper bounds of 2.4 ug/m^3^ and 5.9 ug/m^3^, respectively (Burnett et al. 2014). The dose-response function for the six disease end-points are presented in Figure 1. This graph presents the average value of relative risks at each concentration value, calculated for 1000 iterations of the four IER parameters *ϑ, ω, δ, x* reported by Burnett (2021).

**Figure 1:**
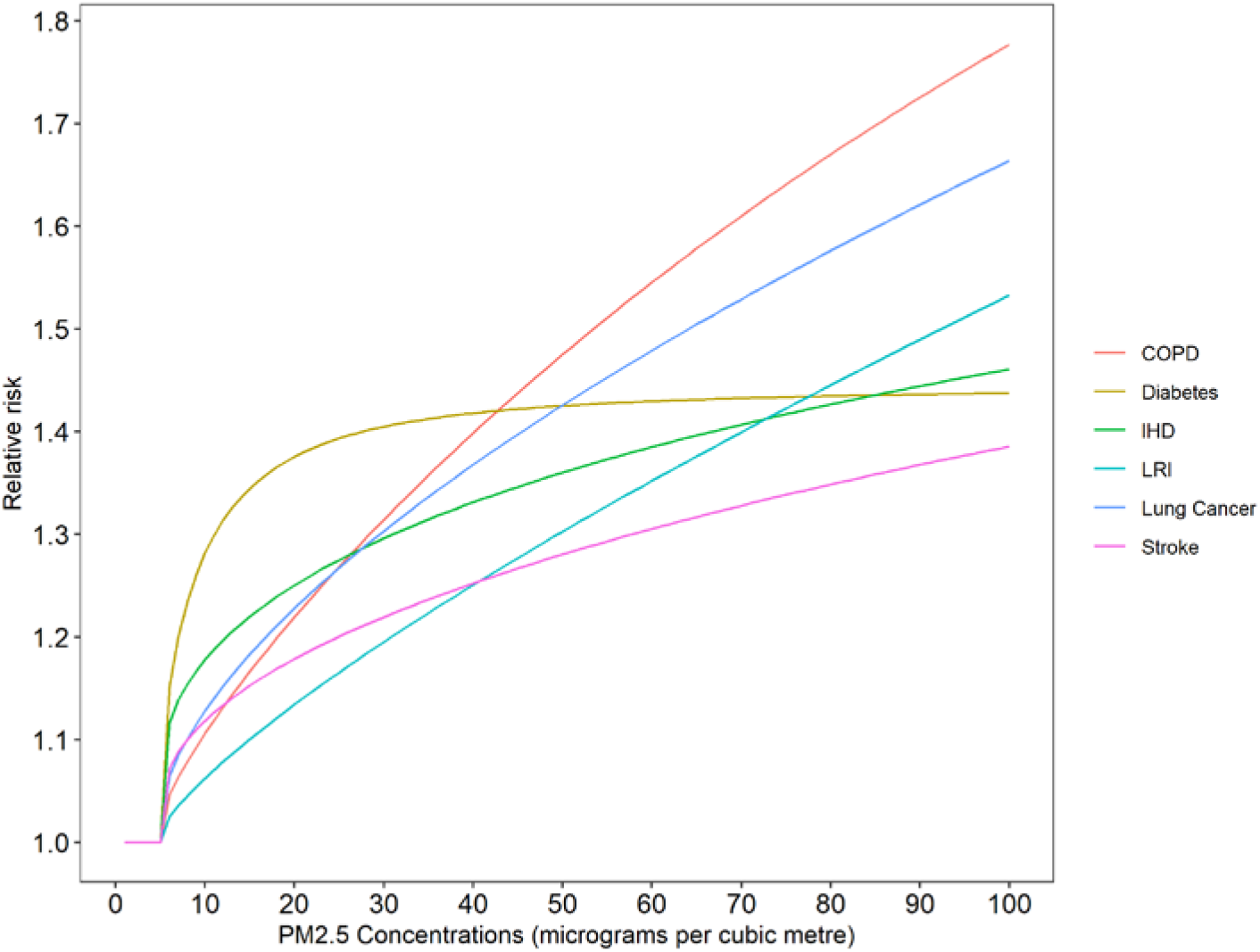
Integrated exposure response function (age group 55-60 years for IHD and stroke)

We used comparative risk assessment approach to estimate the population attributable fraction (PAF) for each city and expressed it as percentage (see equation 4). We defined counterfactual PM_2.5*cf*_ as the concentrations achieved after the complete removal of traffic emissions.

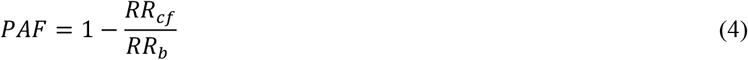

where *RR*_*cf*_ and *RR*_*b*_ are, respectively, the relative risk values (see equation 3) for the concentrations of PM_2.5*cf*_ and PM_2.5*b*_; the latter stands for overall PM_2.5_ concentrations at the baseline. For each city, we calculated PM_2.5*b*_ as the mean value of the posterior densities of overall PM_2.5_ concentrations. We calculated PM_2.5*cf*_ as the difference between the mean values of the posterior densities of overall and traffic-related PM_2.5_. Lastly, using the pair of these concentrations, for each city, we calculated PAF for each of the 1000 iterations of the parameters in the IER functions, and we present mean PAF of those iterations.

## 3. Results

The results relating to the estimated posterior densities of PM_2.5_ for the cities included in our study are reported in the supplementary material (Tables C-E). Note that to investigate robustness of the multivariate normal density assumption in (1), we used a scale mixing approach that can address skewness in the data; this confirmed the suitability of our assumption.

### 3.1. Posterior densities of the meta-regression coefficients

As mentioned previously, we considered variables available in the WHO database to develop our models, detailed in Section 2.3. Through an exploratory data analysis, we noticed a relatively significant difference between measurements reported by studies published before 2005 and those published after 2005 with respect to traffic-related PM_2.5_ concentrations. This is in accordance with Heydari et al. (2020). We therefore created a categorical variable for publication year with the aim of capturing the above difference in the model. Also, we created several categorical variables to include the location of studies in the models, considering different possibilities and combinations of regions. The final location variable had four categories: (1) North America, Oceania or Japan; (2) North Western or Western Europe; (3) rest of Europe; and (4) rest of the world. Japan being a high-income country and having PM_2.5_ concentrations similar to North America and Oceania was included in the first category mentioned above. Table 3 reports the posterior summary of the estimated regression coefficients.

**Table 3.** Meta-regression estimation results

| Variable | Mean | Std. Dev. | 95% Bayesian interval |  |
| --- | --- | --- | --- | --- |
| ***Traffic-related $PM_{2.5}$*** |  |  |  |  |
| Ln(city population) | 0.21 | 0.03 | 0.14 | 0.27 |
| Latitude (divided by 10) | 0.06 | 0.03 | 0.01 | 0.11 |
| Sea salt contribution reported<sup>1</sup> | -0.29 | 0.11 | -0.51 | -0.07 |
| Study published after 2005<sup>2</sup> | -0.36 | 0.11 | -0.57 | -0.14 |
| Study was conducted in North America, Oceania or Japan<sup>3</sup> | -1.25 | 0.13 | -1.50 | -0.99 |
| Study was conducted in North Western or Western Europe<sup>3</sup> | -1.69 | 0.20 | -2.08 | -1.30 |
| Constant | 2.54 | 0.16 | 2.23 | 2.86 |
| ***Overall $PM_{2.5}$*** |  |  |  |  |
| Ln(city population) | 0.08 | 0.03 | 0.02 | 0.13 |
| Latitude (divided by 10) | 0.13 | 0.02 | 0.08 | 0.17 |
| Sea salt contribution reported<sup>1</sup> | -0.33 | 0.08 | -0.50 | -0.17 |
| Study was conducted in North America, Oceania or Japan<sup>3</sup> | -1.11 | 0.11 | -1.33 | -0.89 |
| Study was conducted in North Western or Western Europe<sup>3</sup> | -1.49 | 0.16 | -1.81 | -1.17 |
| Study was conducted in the rest of Europe<sup>3</sup> | -0.41 | 0.11 | -0.63 | -0.19 |
| Constant | 3.63 | 0.09 | 3.46 | 3.81 |

<sup>1</sup> Group of studies that do not report sea salt contribution is the reference group
<sup>2</sup> Rest of the world is the reference group
<sup>3</sup> Group of studies conducted on or before 2005 is the reference group

### 3.2. Correlation between overall PM_2.5_ and traffic-related PM_2.5_

Our modelling approach allowed us to estimate the magnitude of correlation between overall PM_2.5_ and traffic-related PM_2.5_ across a sample of 117 cities worldwide. The mean (standard deviation) of the correlation is 0.63 (0.05), with a 95% Bayesian interval varying from 0.54 to 0.72. This indicates that the correlation between overall and traffic-related PM_2.5_ is statistically important and that traffic-related PM_2.5_ and overall PM_2.5_ are highly correlated.

### 3.3. Health benefit potential of reducing traffic emissions

Figure 2 presents the probabilities of PM_2.5_ concentrations exceeding the WHO-recommended concentration of PM_2.5_ if all traffic emissions are removed in the cities under investigation. This probability accounts for both PM_2.5_ concentrations as well as their associated uncertainties, both of which are represented by posterior distribution of concentrations estimated using our meta-analysis. The graph shows that, when the overall concentration is beyond 20 ug/m^3^, the probability of exceeding the WHO-recommended concentration of PM_2.5_ remains greater than 70 percent.

**Figure 2:**
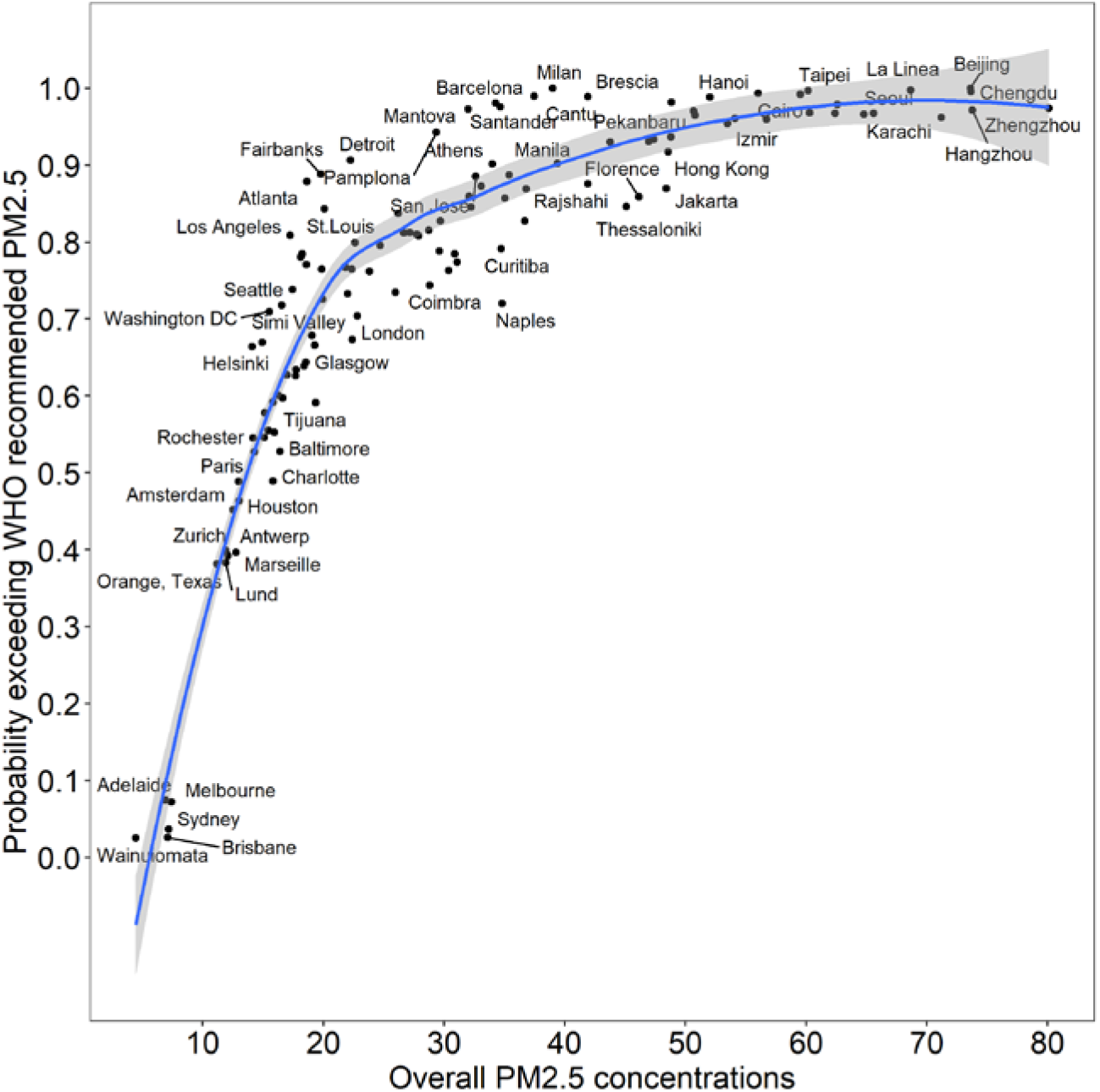
Probability of exceeding WHO-recommended PM_2.5_ if traffic emissions are removed

Figure 3 presents percent reduction in the premature mortality due to four disease end-points (COPD, IHD, Lung Cancer and Diabetes) for a counterfactual scenario of complete removal of traffic-related PM_2.5_ concentrations. Note that Stroke and LRI have similar shaped curves as IHD and Lung Cancer, respectively, and their results are not shown here. According to these graphs, the benefits of prevented mortality increase up to a point (30-40 ug/m^3^) for COPD, IHD and Lung Cancer, at which point there are large variations, and then flatten off. The flattening is most prominent for COPD and Lung Cancer, and less so for IHD. The Diabetes-related mortality reduction show that the largest benefits are limited to concentrations below 25 ug/m^3^, at which point there is large variation across the cities. Above this concentration, the benefits in terms of reduced mortality become progressively smaller.

**Figure 3:**
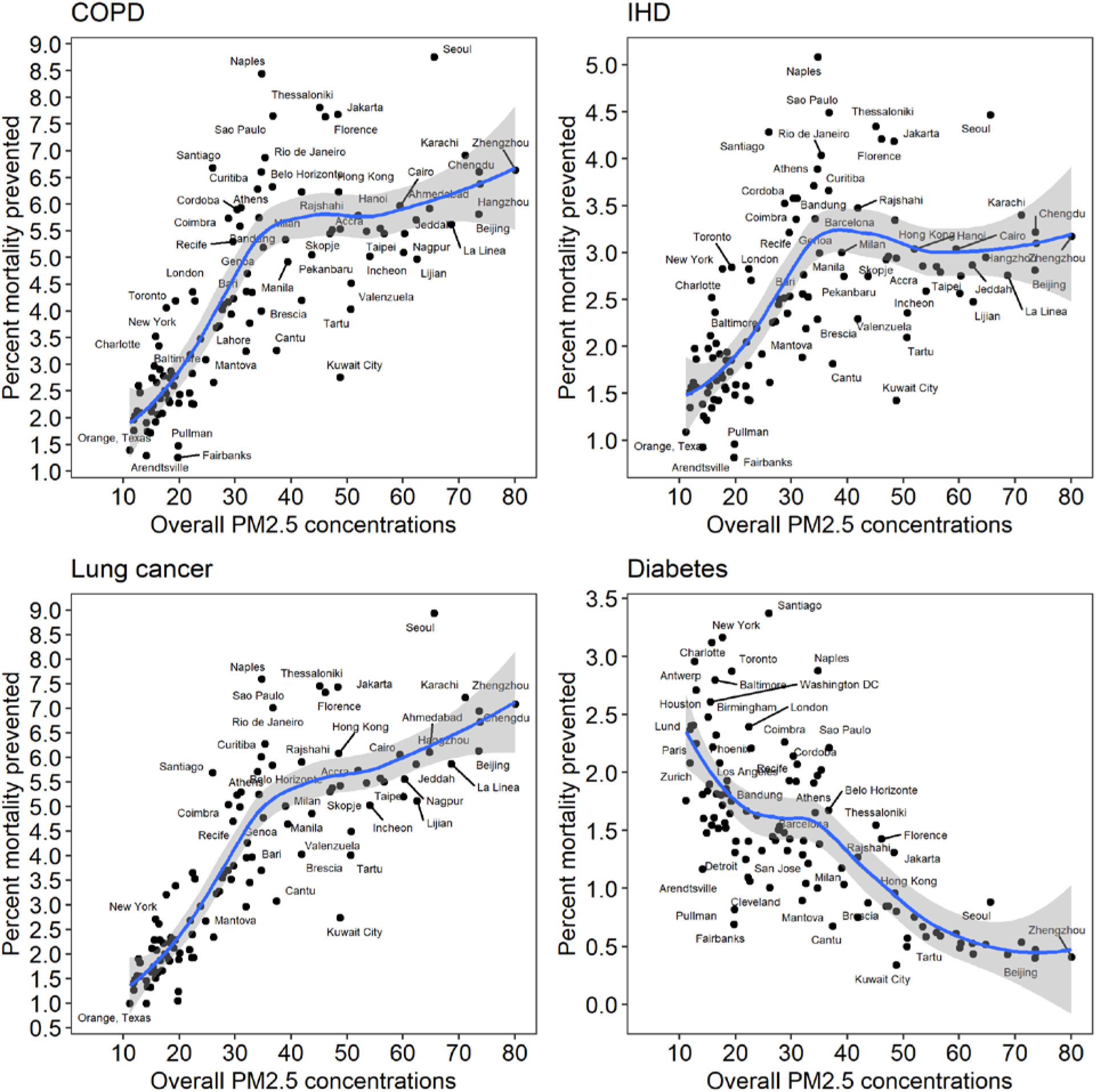
Percentage reduction in mortality resulting from total removal of traffic emissions (for COPD and IHD, age group= 50-55 years)

## 4. Discussion

### 4.1. Statement of principal findings

We used the WHO database of source apportionment studies to estimate overall, traffic-related, and traffic-excluded PM_2.5_ concentrations using a Bayesian meta-regression approach. For the posterior distributions of the expected concentration of non-traffic-related PM_2.5_, we estimated the probability that concentrations remained higher than the WHO-recommended annual concentration of PM_2.5_ if traffic-related emissions were removed completely. We found that this probability rises steeply up to 30 ug/m^3^, and then remains greater than 70 percent. In other words, for the cities with overall concentrations greater than 30 ug/m^3^, the removal of traffic-related emissions are highly unlikely to reduce concentration levels down to the WHO-recommended PM_2.5_ annual guideline.

We estimated the percentage reduction in premature mortality due to four disease end-points (COPD, IHD, Lung Cancer and Diabetes) if all traffic-related emissions are removed. We used non-linear IER functions along with the comparative risk assessment approach to estimate population attributable fraction corresponding to this reduction in concentrations. We found that for COPD, IHD and Lung Cancer, the percent reduction in mortality increases up to 30-40 ug/m^3^, and at higher concentrations, it flattens off, showing diminishing returns. The flattening is far more prominent in IHD than in COPD and Lung Cancer. In Figure 1, IER for COPD and Lung Cancer have steeper increase in relative risk than IHD. In case of Diabetes, the pattern is opposite to that of the other three outcomes. With increasing concentrations, there are negative returns in the reduction of premature mortality. This is expected given that Diabetes is the only disease outcome that has a prominently flat IER function after a steep jump up to 20 ug/m^3^. Mathematically, a flat function implies that the ratio of relative risks in equation 3 approaches unity at higher concentrations.

### 4.2. Strengths and weaknesses of the study

We employed a multivariate meta-regression approach to estimate the expected concentrations of PM_2.5_ of different types (in terms of source) for 117 cities in various regions worldwide. Note that compared to relaying on information from one study only, conclusions from a meta-analytic approach are more reliable for evidence-based policy making. Further, we modelled traffic-related and overall PM_2.5_ concentrations jointly; therefore, our estimates have superior statistical properties. This is due to the fact that data points borrow strength from other related data points (Jackson et al., 2011). Also, a joint analysis allowed us to investigate the magnitude of correlation between traffic related PM_2.5_ and overall PM_2.5_ based on a systematic approach, providing further insight into the relationship between the two types of concentrations. Understanding this correlation is quite interesting as it allows estimating the range of one type of concentration from the other one when both pieces of information are not available for a given location. Note that our meta-regression, being developed under the Bayesian paradigm, accounted for uncertainties in both regression parameter estimates and predicted values fully.

One of the limitations of our study is that we have only estimated reduction in mortality due to different diseases as percentage of baseline health burden. In terms of mortality rates (e.g. deaths per 100,000 persons), these percentages could translate into highly divergent values. This is because, in certain countries that have much greater proportion of older adults, baseline incidence rates of mortality from the cardiovascular, respiratory, and metabolic diseases are much greater than countries with much younger population. Also, LMICs have much greater incidence rates of mortality than HICs. Secondly, many city estimates used in our analysis are more than a decade old or even older. As a result, our study does not represent the latest situation of most of the cities. However, our study provides an analytical approach that can be readily updated as new data (other studies) becomes available.

### 4.3. Meaning of the study: possible mechanisms and implications for policymakers

We found that only for a handful of cities could reduction in traffic emissions result in achieving the WHO-recommended guideline for annual PM_2.5_ concentrations. We used the WHO guideline for the purpose of illustration, as for many settings across the world, this is a highly ambitious scenario. The method presented here can be applied for more realistic targets of pollution concentrations and could include reductions across multiple sectors. Using a stochastic approach, we can make a probabilistic judgement of the impact that policies will have. Our approach, being developed under the Bayesian framework, can be used to estimate updated probabilities as more information is added or updated information is added for the same cities. The health benefits that we presented are also for a highly ambitious scenario, in which all of traffic emissions are removed. However, due to non-linearity in IER functions, we found that health benefits are not proportionally as large. While traffic as a polluting sector gains a lot of attention due to visibility of its sources (i.e. vehicles), our results imply that cities with high concentrations of pollution need a multi-sectoral framework to reduce anthropogenic emissions. This will not only help clean air much faster, it will also make investments more cost-effective as concentrations near the steeper part of the curve.

PM_2.5_ emissions from vehicles are largely proportional to sulphur content in the fuel, and cleaner fuel with lower sulphur content can significantly reduce traffic emissions (CCAC, 2016; Goel & Guttikunda, 2015). While Europe and North America implemented use of low-and ultra-low-sulphur diesel (less than 50 ppm and less than 15 ppm sulphur content, respectively) in late 2000s (EEA, 2016), the progress in many of the low-and-middle income countries has been much slower (CCAC, 2016). A timeline of sulphur concentrations in diesel for 2013-2020 period across the world (UNEP/CCAC, 2018) shows that many countries are gradually progressing towards the use of low-sulphur fuel. However, more than half of the world’s countries are still using high-sulphur fuels. These are mainly low-and middle-income countries spread across Latin America, the Caribbean, Africa, the Middle East, and Asia-Pacific (CCAC, 2016). It is only with low-sulphur fuels that vehicles with stricter emission standards can be effective. Thus, accelerating the desulphurisation of fuel and adoption of cleaner vehicle standards in large parts across the world has large potential to prevent health burden attributed to traffic.

### 4.4. Unanswered questions and future research

As highlighted in the limitations, the underlying dataset of cities (WHO, 2015) reporting source apportionment has studies that were done a decade or longer ago. It is likely that since this database was created about five years ago, an updated review of source-apportionment studies could greatly improve our understanding of different sources of pollution in cities. The results on health impacts presented here are based on IER functions that were used in GBD 2017. The different types of risk functions for PM_2.5_ have been shown to have significant impact on their respective estimates of disease burden (Evangelopoulos et al., 2020). As evidence improves on different health outcomes as well as from settings with high concentrations of pollution, we could expect changes in these functions and their respective health impact estimates.

## Data Availability

The data can be obtained from the following link

http://www.who.int/quantifying_ehimpacts/global/source_apport/

