## Supplementary material for "Potential Health Benefits of Eliminating Traffic Emissions in Urban Areas"

### Table A. List of studies included in our meta-analysis

| List of studies | Reference year | City | Country | Region |
| --- | --- | --- | --- | --- |
| AIRPARIF/LSCE220 | 2011 | Paris | France | Western Europe |
| AIRPARIF/LSCE220 | 2011 | Paris | France | Western Europe |
| Aldabe208 | 2011 | Pamplona | Spain | Southwestern Europe |
| Aldabe208 | 2011 | Pamplona | Spain | Southwestern Europe |
| Alolayan53 | 2013 | Kuwait City | Kuwait | Middle East |
| Amato128–131 | 2014 | Alcala de Guadaira | Spain | Southwestern Europe |
| Amato128–131 | 2009 | Barcelona | Spain | Southwestern Europe |
| Amato128–131 | 2014 | Barcelona | Spain | Southwestern Europe |
| Andrade1 | 2012 | Belo Horizonte | Brazil | South/Central America |
| Andrade1 | 2012 | Curitiba | Brazil | South/Central America |
| Andrade1 | 2012 | Porto Alegre | Brazil | South/Central America |
| Andrade1 | 2012 | Recife | Brazil | South/Central America |
| Andrade1 | 2012 | Rio de Janeiro | Brazil | South/Central America |
| Andrade1 | 2012 | Sao Paulo | Brazil | South/Central America |
| Arruti135 | 2011 | Santander | Spain | Southwestern Europe |
| Begum34 | 2005 | Rajshahi | Bangladesh | Southern Asia |
| Begum86 | 2005 | Washington DC | USA | North America |
| Boman56 | 2012 | Cairo | Egypt | Middle East |
| Bruno138 | 2008 | Bari | Italy | Southwestern Europe |
| Chan57,58 | 2008 | Adelaide | Australia | Oceana/Japan |
| Chan57,58 | 2008 | Brisbane | Australia | Oceana/Japan |
| Chan57,58 | 2011 | Brisbane | Australia | Oceana/Japan |
| Chan57,58 | 2008 | Melbourne | Australia | Oceana/Japan |
| Chan57,58 | 2008 | Sydney | Australia | Oceana/Japan |
| Cheng14 | 2014 | Hong Kong | China | East Asia |
| Chiou89–91 | 2008 | Houston | USA | North America |
| Chiou89–91 | 2008 | Orange, Texas | USA | North America |
| Choi45 | 2013 | Incheon | Korea | East Asia |
| Cohen59 | 2011 | Sydney | Australia | Oceana/Japan |
| Cohen70,71 | 2010 | Hanoi | Vietnam | Southeastern Asia |
| Cohen70,71 | 2008 | Manila | Philippines | Southeastern Asia |
| Coutant92,93 | 2002 | Arendtsville | USA | North America |
| Coutant92,93 | 2003 | Birminghamusa | USA | North America |
| Coutant92,93 | 2003 | Charlotte | USA | North America |
| Coutant92,93 | 2003 | Houston | USA | North America |
| Coutant92,93 | 2003 | Indianapolis | USA | North America |
| Coutant92,93 | 2003 | Milwaukee | USA | North America |
| Coutant92,93 | 2003 | New York | USA | North America |
| Coutant92,93 | 2003 | St.Louis | USA | North America |
| Coutant92,93 | 2002 | Washington DC | USA | North America |
| Coutant92,93 | 2003 | Washington DC | USA | North America |
| D'Alessandro145 | 2003 | Florence | Italy | Southwestern Europe |
| D'Alessandro145 | 2003 | Naples | Italy | Southwestern Europe |
| Davy60 | 2014 | Wainuiomata | New Zeland | Oceana/Japan |
| DEFRA146 | 2005 | Belfast | UK | Northwestern Europe |
| DEFRA146 | 2005 | Birmingham | UK | Northwestern Europe |
| DEFRA146 | 2005 | Glasgow | UK | Northwestern Europe |
| DEFRA146 | 2005 | London | UK | Northwestern Europe |
| DEFRA146 | 2005 | Manchester | UK | Northwestern Europe |
| DEFRA146 | 2005 | Rochester | UK | Northwestern Europe |
| El Haddad147 | 2011 | Marseille | France | Western Europe |
| ESMAP report4 | 2011 | Cairo | Egypt | Middle East |
| ESMAP report4 | 2011 | Santiago | Chile | South/Central America |
| ESMAP report4 | 2011 | Sao Paulo | Brazil | South/Central America |
| Friend61 | 2011 | Brisbane | Australia | Oceana/Japan |
| Geng16 | 2013 | Zhengzhou | China | East Asia |
| Godoy5 | 2009 | Rio de Janeiro | Brazil | South/Central America |
| Green94 | 2013 | Las Vegas | USA | North America |
| Gummeneni35 | 2011 | Hyderabad | India | Southern Asia |
| Guttikunda36 | 2012 | Hyderabad | India | Southern Asia |
| Hai72 | 2013 | Hanoi | Vietnam | Southeastern Asia |
| Hammond95 | 2008 | Detroit | USA | North America |
| Harrison218 | 1997 | Birmingham | UK | Northwestern Europe |
| Harrison218 | 1997 | Coimbra | Portugal | Southwestern Europe |
| Hasheminassab222 | 2014 | Los Angeles | USA | North America |
| Hasheminassab223 | 2014 | Bakersfield | USA | North America |
| Hasheminassab223 | 2014 | Fresno | USA | North America |
| Hasheminassab223 | 2014 | Los Angeles | USA | North America |
| Hasheminassab223 | 2014 | Sacramento | USA | North America |
| Hasheminassab223 | 2014 | San Joseusa | USA | North America |
| Hasheminassab223 | 2014 | Simi Valley | USA | North America |
| Heon-Jeong6,7 | 2011 | Edmonton | Canada | North America |
| Heon-Jeong6,7 | 2011 | Halifax | Canada | North America |
| Heon-Jeong6,7 | 2011 | Montreal | Canada | North America |
| Heon-Jeong6,7 | 2011 | Windsor | Canada | North America |
| Iijima44 | 2008 | Tokyo | Japan | Oceana/Japan |
| Ilacqua157 | 2007 | Athens | Greece | Southwestern Europe |
| Ilacqua157 | 2007 | Helsinki | Finland | Northwestern Europe |
| Jimenez96 | 2006 | Pullman | USA | North America |
| Jorquera63–65 | 2012 | Santiago | Chile | South/Central America |
| Karanasiou158–160 | 2009 | Athens | Greece | Southwestern Europe |
| Khodeir54 | 2012 | Jeddah | Saudi Arabia | Middle East |
| Kim225 | 2010 | Los Angeles | USA | North America |
| Kim228 | 2007 | Los Angeles | USA | North America |
| Kim228 | 2007 | Simi Valley | USA | North America |
| Kim97–103 | 2002 | Atlanta | USA | North America |
| Kim97–103 | 2003 | Atlanta | USA | North America |
| Kim97–103 | 2003 | Atlanta | USA | North America |
| Kim97–103 | 2003 | Washington DC | USA | North America |
| Kovacevik10 | 2011 | Skopje | Macedonia | Central and Eastern Europe |
| Larsen162 | 2010 | Brescia | Italy | Southwestern Europe |
| Larsen162 | 2010 | Brescia | Italy | Southwestern Europe |
| Larsen162 | 2010 | Cantu | Italy | Southwestern Europe |
| Larsen162 | 2010 | Cantu | Italy | Southwestern Europe |
| Larsen162 | 2010 | Mantova | Italy | Southwestern Europe |
| Larsen162 | 2010 | Mantova | Italy | Southwestern Europe |
| Larsen162 | 2010 | Milan | Italy | Southwestern Europe |
| Larsen162 | 2010 | Milan | Italy | Southwestern Europe |
| Larsen162 | 2010 | Milan | Italy | Southwestern Europe |
| Larsen162 | 2010 | Milan | Italy | Southwestern Europe |
| Lee226 | 2006 | St.Louis | USA | North America |
| Lee8 | 2003 | Toronto | Canada | North America |
| Lee8,104 | 2008 | Torontousa | USA | North America |
| Lestari74 | 2009 | Bandung | Indonesia | Southeastern Asia |
| Lewis105 | 2002 | Phoenix | USA | North America |
| Li106 | 2004 | New York | USA | North America |
| Liang20 | 2013 | Taipei | Taiwan | East Asia |
| Liang20 | 2013 | Taipei | Taiwan | East Asia |
| Liu107 | 2006 | Atlanta | USA | North America |
| Liu107 | 2006 | Birminghamusa | USA | North America |
| Liu21 | 2014 | Hangzhou | China | East Asia |
| Lodhi37 | 2009 | Lahore | Pakistan | Southern Asia |
| Lopez66 | 2011 | Cordoba | Argentina | South/Central America |
| Manoli165 | 2002 | Thessaloniki | Greece | Southwestern Europe |
| Mansha38 | 2012 | Karachi | Pakistan | Southern Asia |
| Marcazzan166 | 2003 | Milan | Italy | Southwestern Europe |
| Maykut109 | 2003 | Seattle | USA | North America |
| Maykut109 | 2003 | Seattle | USA | North America |
| Maykut109 | 2003 | Seattle | USA | North America |
| Mazzei170 | 2008 | Genoa | Italy | Southwestern Europe |
| Minguillon50 | 2014 | Tijuana | Mexico | North America |
| Moreno171,172 | 2011 | Santander | Spain | Southwestern Europe |
| Morishita110 | 2006 | Detroit | USA | North America |
| Motallebi220 | 1999 | Sacramento | USA | North America |
| Murillo52 | 2012 | Salamanca | Mexico | North America |
| Murillo67 | 2013 | San Jose | Costa Rica | South/Central America |
| Ogulei111 | 2005 | Baltimore | USA | North America |
| Orru11 | 2010 | Tartu | Estonia | Central and Eastern Europe |
| Pabroa76 | 2011 | Valenzuela | Philippines | Southeastern Asia |
| Pandolfi175,176 | 2010 | Algeciras | Spain | Southwestern Europe |
| Pandolfi175,176 | 2010 | La Linea | Spain | Southwestern Europe |
| Pandolfi175,176 | 2010 | Los Barrios | Spain | Southwestern Europe |
| Park48 | 2005 | Seoul | Korea | East Asia |
| Park48 | 2005 | Seoul | Korea | East Asia |
| Perrone214 | 2011 | Milan | Italy | Southwestern Europe |
| Perrone214 | 2011 | Milan | Italy | Southwestern Europe |
| Perrone214 | 2011 | Milan | Italy | Southwestern Europe |
| Perrone214 | 2011 | Milan | Italy | Southwestern Europe |
| Perrone214 | 2011 | Milan | Italy | Southwestern Europe |
| Pipalatkar40 | 2014 | Nagpur | India | Southern Asia |
| Querol178–180 | 2002 | Barcelona | Spain | Southwestern Europe |
| Rahman77 | 2011 | Kuala Lumpur | Malaysia | Southeastern Asia |
| Raja41 | 2010 | Lahore | Pakistan | Southern Asia |
| Richard210 | 2011 | Zurich | Switzerland | Western Europe |
| Sanchez184,185 | 2007 | Huelva | Spain | Southwestern Europe |
| Santoso78 | 2013 | Jakarta | Indonesia | Southeastern Asia |
| See79 | 2007 | Pekanbaru | Indonesia | Southeastern Asia |
| Seneviratne42 | 2011 | Colombo | Sri Lanka | Southern Asia |
| Song115 | 2001 | Washington DC | USA | North America |
| Song23 | 2006 | Beijing | China | East Asia |
| Song23 | 2006 | Beijing | China | East Asia |
| Song23 | 2006 | Beijing | China | East Asia |
| Song23 | 2006 | Beijing | China | East Asia |
| Song23 | 2007 | Beijing | China | East Asia |
| Sowka12 | 2012 | Wrokalw | Poland | Central and Eastern Europe |
| Sudheer43 | 2012 | Ahmedabad | India | Southern Asia |
| Swietlicki219 | 1996 | Lund | Sweden | Northwestern Europe |
| Tahir75 | 2013 | Kuala Terengganu | Malaysia | Southeastern Asia |
| Tao24 | 2013 | Chengdu | China | East Asia |
| Tian25 | 2013 | Chengdu | China | East Asia |
| Vallius189,190 | 2005 | Amsterdam | The Netherlands | Western Europe |
| Vallius189,190 | 2005 | Erfurt | Germany | Western Europe |
| Vallius189,190 | 2003 | Helsinki | Finland | Northwestern Europe |
| Vallius189,190 | 2005 | Helsinki | Finland | Northwestern Europe |
| Van Borm217 | 1987 | Antwerp | Belgium | Western Europe |
| Viana191–194 | 2007 | Albacete | Spain | Southwestern Europe |
| Viana191–194 | 2007 | Barcelona | Spain | Southwestern Europe |
| Viana191–194 | 2007 | Oviedo | Spain | Southwestern Europe |
| Wang117,118 | 2014 | Fairbanks | USA | North America |
| Wang117,118 | 2013 | San Joseusa | USA | North America |
| Wang26 | 2007 | Beijing | China | East Asia |
| Ward119 | 2012 | Fairbanks | USA | North America |
| Yatkin84,85 | 2008 | Izmir | Turkey | Middle East |
| Yli-Tuomi200,201 | 2007 | Amsterdam | The Netherlands | Western Europe |
| Yli-Tuomi200,201 | 2007 | Helsinki | Finland | Northwestern Europe |
| Yu29 | 2013 | Beijing | China | East Asia |
| Yuling120 | 2010 | Dallas | USA | North America |
| Zhang32 | 2013 | Lijian | China | East Asia |
| Zhao229 | 2006 | Indianapolis | USA | North America |
| Zhou122,123 | 2013 | Accra | Ghana | Africa |
| Zhou122,123 | 2004 | Pittsburg | USA | North America |
| Zhou230 | 2009 | Cleveland | USA | North America |
| ^(1)^ For a complete list of studies, see the World Health Organisation's Source Apportionment Database for PM10 and PM2.5 Updated to August 2014. http://www.who.int/quantifying_ehimpacts/global/source_apport/. | | | | |

**Table B. Country grouping**

| **Region** | **Countries** |
| --- | --- |
| Africa | Ghana |
| Central and Eastern Europe | Estonia, Macedonia, Poland |
| East Asia | China, Korea, Taiwan |
| Middle East | Egypt, Kuwait, Saudi Arabia, Turkey |
| North America | Canada, Mexico, USA |
| Northwestern Europe | Finland, Sweden, UK |
| Oceana/Japan | Australia, Japan, New Zeland |
| South/Central America | Argentina, Brazil, Chile, Costa Rica |
| Southeastern Asia | Indonesia, Malaysia, Philippines, Vietnam |
| Southern Asia | Bangladesh, India, Pakistan, Sri Lanka |
| Southwestern Europe | Greece, Italy, Portugal, Spain |
| Western Europe | Belgium, France, Germany, Switzerland, The Netherlands |

**Table C. Estimated expected posterior summaries of traffic-related PM_2.5_**

|  |  |  |  | 95% Bayesian interval | |
| --- | --- | --- | --- | --- | --- |
| City | Mean | St.Dev. | Median | 2.50% | 97.50% |
| Accra | 13.37 | 11.6 | 10.11 | 2.353 | 43.15 |
| Adelaide | 2.213 | 1.994 | 1.663 | 0.3806 | 7.379 |
| Ahmedabad | 17.17 | 14.86 | 12.99 | 3.101 | 56.36 |
| Albacete | 7.547 | 6.548 | 5.683 | 1.353 | 24.54 |
| Alcala de Guadaira | 6.304 | 5.382 | 4.797 | 1.122 | 20.63 |
| Algeciras | 6.867 | 5.825 | 5.245 | 1.202 | 22.18 |
| Amsterdam | 2.162 | 1.265 | 1.872 | 0.6348 | 5.443 |
| Antwerp | 2.642 | 2.361 | 1.979 | 0.4641 | 8.915 |
| Arendtsville | 1.472 | 1.335 | 1.098 | 0.2394 | 4.934 |
| Athens | 11.85 | 6.764 | 10.29 | 3.609 | 29.02 |
| Atlanta | 3.683 | 1.5 | 3.402 | 1.592 | 7.35 |
| Bakersfield | 2.489 | 2.113 | 1.888 | 0.4417 | 8.054 |
| Baltimore | 4.043 | 3.566 | 3.044 | 0.7213 | 13.22 |
| Bandung | 9.993 | 8.565 | 7.588 | 1.788 | 32.95 |
| Barcelona | 10.99 | 4.31 | 10.24 | 4.808 | 21.36 |
| Bari | 8.773 | 7.553 | 6.698 | 1.58 | 28.49 |
| Beijing | 18.39 | 5.741 | 17.53 | 9.636 | 31.8 |
| Belfast | 3.198 | 2.803 | 2.412 | 0.5515 | 10.44 |
| Belo Horizonte | 12.55 | 10.89 | 9.433 | 2.161 | 41.73 |
| Birmingham | 3.18 | 1.88 | 2.736 | 0.9431 | 8.036 |
| Birminghamusa | 3.058 | 1.738 | 2.653 | 0.9502 | 7.521 |
| Brescia | 9.358 | 5.402 | 8.1 | 2.838 | 23.15 |
| Brisbane | 2.105 | 1.026 | 1.891 | 0.7695 | 4.695 |
| Cairo | 16.36 | 9.458 | 14.19 | 5.017 | 40.41 |
| Cantu | 6.846 | 4.048 | 5.907 | 2.037 | 17.24 |
| Charlotte | 4.128 | 3.524 | 3.117 | 0.7231 | 13.67 |
| Chengdu | 20.74 | 11.94 | 18.02 | 6.306 | 51.15 |
| Cleveland | 3.482 | 3.038 | 2.623 | 0.6037 | 11.54 |
| Coimbra | 9.768 | 8.625 | 7.334 | 1.69 | 32.7 |
| Colombo | 8.299 | 7.193 | 6.295 | 1.476 | 26.81 |
| Cordoba | 10.37 | 9.039 | 7.833 | 1.786 | 34.24 |
| Curitiba | 12.57 | 11.03 | 9.431 | 2.188 | 41.36 |
| Dallas | 4.274 | 3.721 | 3.24 | 0.7386 | 14.03 |
| Detroit | 3.45 | 1.972 | 2.994 | 1.065 | 8.496 |
| Edmonton | 3.22 | 2.841 | 2.411 | 0.5546 | 10.7 |
| Erfurt | 2.949 | 2.638 | 2.226 | 0.4891 | 9.766 |
| Fairbanks | 1.814 | 1.061 | 1.563 | 0.5423 | 4.54 |
| Florence | 17.3 | 15.23 | 13.07 | 3.014 | 56.64 |
| Fresno | 2.65 | 2.321 | 2.022 | 0.4629 | 8.501 |
| Genoa | 10.14 | 8.737 | 7.672 | 1.797 | 33.22 |
| Glasgow | 3.82 | 3.439 | 2.841 | 0.6526 | 12.95 |
| Halifax | 2.655 | 2.31 | 2.008 | 0.4524 | 8.655 |
| Hangzhou | 20.1 | 17.61 | 15.3 | 3.539 | 65.63 |
| Hanoi | 14.56 | 8.383 | 12.68 | 4.451 | 36.03 |
| Helsinki | 2.131 | 0.9146 | 1.959 | 0.8902 | 4.392 |
| Hong Kong | 14.89 | 13.01 | 11.2 | 2.603 | 49.75 |
| Houston | 2.556 | 1.448 | 2.224 | 0.7808 | 6.185 |
| Huelva | 7.271 | 6.369 | 5.506 | 1.288 | 23.87 |
| Hyderabad | 14.67 | 8.273 | 12.71 | 4.551 | 35.87 |
| Incheon | 13.06 | 11.31 | 9.946 | 2.285 | 42.79 |
| Indianapolis | 3.669 | 2.142 | 3.177 | 1.124 | 9.238 |
| Izmir | 14.55 | 13.12 | 10.95 | 2.518 | 47.92 |
| Jakarta | 17.97 | 15.56 | 13.56 | 3.236 | 59.29 |
| Jeddah | 16.2 | 14.01 | 12.18 | 2.917 | 52.05 |
| Karachi | 21.14 | 18.02 | 16.18 | 3.657 | 67.82 |
| Kuala Lumpur | 13.09 | 11.27 | 9.875 | 2.331 | 42.85 |
| Kuala Terengganu | 4.476 | 3.952 | 3.359 | 0.7478 | 14.78 |
| Kuwait City | 6.875 | 6.151 | 5.171 | 1.164 | 22.31 |
| La Linea | 17.03 | 9.694 | 14.84 | 5.217 | 42.13 |
| Lahore | 6.174 | 5.435 | 4.64 | 1.066 | 20.45 |
| Las Vegas | 3.696 | 3.203 | 2.806 | 0.6515 | 12.16 |
| Lijian | 14.23 | 12.19 | 10.78 | 2.542 | 45.86 |
| London | 6.411 | 5.855 | 4.761 | 1.08 | 21.29 |
| Los Angeles | 3.535 | 1.431 | 3.279 | 1.531 | 7.041 |
| Los Barrios | 4.969 | 4.307 | 3.774 | 0.8767 | 16.38 |
| Lund | 1.925 | 1.717 | 1.444 | 0.3286 | 6.412 |
| Manchester | 3.647 | 3.272 | 2.723 | 0.6121 | 12.25 |
| Manila | 10.42 | 8.702 | 8.037 | 1.815 | 33.27 |
| Mantova | 6.156 | 3.606 | 5.317 | 1.856 | 15.21 |
| Marseille | 2.009 | 1.8 | 1.494 | 0.3394 | 6.801 |
| Melbourne | 2.869 | 2.587 | 2.14 | 0.4739 | 9.559 |
| Milan | 11.16 | 2.835 | 10.8 | 6.686 | 17.73 |
| Milwaukee | 5.42 | 4.621 | 4.144 | 0.952 | 17.48 |
| Montreal | 3.546 | 3.028 | 2.688 | 0.6187 | 11.59 |
| Nagpur | 15.16 | 13.19 | 11.49 | 2.663 | 49.41 |
| Naples | 15.62 | 13.59 | 11.78 | 2.732 | 50.37 |
| New York | 5.092 | 2.966 | 4.397 | 1.532 | 12.57 |
| Orange, Texas | 1.324 | 1.142 | 1.01 | 0.2237 | 4.32 |
| Oviedo | 8.158 | 7.03 | 6.221 | 1.453 | 26.5 |
| Pamplona | 6.989 | 4.051 | 6.052 | 2.134 | 17.5 |
| Paris | 2.187 | 1.31 | 1.872 | 0.6387 | 5.567 |
| Pekanbaru | 11.44 | 9.678 | 8.737 | 2.021 | 37.23 |
| Phoenix | 6.264 | 5.48 | 4.757 | 1.095 | 20.47 |
| Pittsburg | 4.74 | 4.113 | 3.581 | 0.8323 | 15.46 |
| Porto Alegre | 10.6 | 9.241 | 7.976 | 1.83 | 34.89 |
| Pullman | 2.122 | 1.845 | 1.592 | 0.3629 | 6.904 |
| Rajshahi | 13.51 | 11.56 | 10.23 | 2.402 | 43.21 |
| Recife | 9.263 | 8.258 | 6.973 | 1.607 | 30.3 |
| Rio de Janeiro | 13.19 | 7.671 | 11.41 | 3.987 | 32.94 |
| Rochester | 1.977 | 1.757 | 1.47 | 0.3342 | 6.673 |
| Sacramento | 3.221 | 1.846 | 2.794 | 0.9818 | 7.908 |
| Salamanca | 2.548 | 2.219 | 1.919 | 0.4459 | 8.227 |
| San Jose | 7.201 | 6.321 | 5.432 | 1.268 | 23.69 |
| San Joseusa | 3.095 | 1.783 | 2.682 | 0.9456 | 7.711 |
| Santander | 7.902 | 4.507 | 6.847 | 2.426 | 19.32 |
| Santiago | 10.41 | 6.184 | 8.964 | 3.051 | 26.18 |
| Sao Paulo | 14.9 | 8.903 | 12.83 | 4.427 | 37.71 |
| Seattle | 3.562 | 1.657 | 3.243 | 1.343 | 7.659 |
| Seoul | 24.81 | 14.47 | 21.45 | 7.439 | 61.52 |
| Simi Valley | 2.012 | 1.149 | 1.747 | 0.6162 | 4.933 |
| Skopje | 12.86 | 11.08 | 9.78 | 2.26 | 41.53 |
| St.Louis | 3.459 | 2.01 | 2.984 | 1.046 | 8.702 |
| Sydney | 2.497 | 1.495 | 2.147 | 0.7269 | 6.308 |
| Taipei | 14.2 | 8.046 | 12.33 | 4.381 | 34.79 |
| Tartu | 10.16 | 8.987 | 7.737 | 1.755 | 33.34 |
| Thessaloniki | 17.4 | 15.32 | 13.07 | 3.047 | 57.1 |
| Tijuana | 3.2 | 2.772 | 2.42 | 0.5552 | 10.35 |
| Tokyo | 7.043 | 6.129 | 5.294 | 1.2 | 23.13 |
| Toronto | 5.591 | 4.935 | 4.218 | 0.9566 | 18.37 |
| Torontousa | 3.826 | 3.272 | 2.901 | 0.6746 | 12.65 |
| Valenzuela | 11.36 | 10.32 | 8.611 | 2.024 | 36.59 |
| Wainuiomata | 0.8184 | 0.7703 | 0.5982 | 0.1291 | 2.814 |
| Washington DC | 3.468 | 1.265 | 3.261 | 1.643 | 6.438 |
| Windsor | 2.341 | 2.064 | 1.758 | 0.4047 | 7.755 |
| Wrokalw | 14.11 | 12.01 | 10.77 | 2.512 | 45.92 |
| Zhengzhou | 22.07 | 19.52 | 16.72 | 3.822 | 72.32 |
| Zurich | 1.727 | 1.582 | 1.283 | 0.2913 | 5.735 |

**Table D. Estimated expected posterior summaries of overall PM_2.5_**

|  |  |  |  | 95% Bayesian interval | |
| --- | --- | --- | --- | --- | --- |
| City | Mean | St.Dev. | Median | 2.50% | 97.50% |
| Accra | 48.79 | 28.71 | 42.13 | 14.24 | 121 |
| Adelaide | 6.904 | 4.247 | 5.869 | 1.975 | 18.02 |
| Ahmedabad | 64.77 | 38.75 | 55.74 | 18.69 | 163.5 |
| Albacete | 29.71 | 18.25 | 25.35 | 8.47 | 76.54 |
| Alcala de Guadaira | 27.17 | 16.23 | 23.41 | 7.964 | 69.32 |
| Algeciras | 27.72 | 16.64 | 23.79 | 8.115 | 70.92 |
| Amsterdam | 12.52 | 5.225 | 11.58 | 5.241 | 25.49 |
| Antwerp | 12.74 | 7.728 | 10.91 | 3.637 | 32.49 |
| Arendtsville | 14.17 | 8.891 | 12.04 | 3.86 | 37.05 |
| Athens | 34 | 14.06 | 31.36 | 14.27 | 68.24 |
| Atlanta | 18.63 | 5.432 | 17.89 | 10.25 | 31.24 |
| Bakersfield | 15.09 | 9.187 | 12.91 | 4.341 | 38.6 |
| Baltimore | 16.38 | 9.761 | 14.13 | 4.808 | 41.11 |
| Bandung | 30.9 | 18.7 | 26.45 | 8.972 | 79.27 |
| Barcelona | 34.27 | 9.989 | 32.88 | 18.9 | 57.29 |
| Bari | 32.23 | 19.2 | 27.79 | 9.412 | 81.12 |
| Beijing | 73.61 | 17 | 71.65 | 45.88 | 112.1 |
| Belfast | 17.74 | 10.78 | 15.22 | 5.042 | 45.71 |
| Belo Horizonte | 36.67 | 21.85 | 31.44 | 10.77 | 92.44 |
| Birmingham | 15.14 | 6.404 | 13.9 | 6.291 | 30.93 |
| Birminghamusa | 18.25 | 7.544 | 16.85 | 7.677 | 36.62 |
| Brescia | 41.92 | 17.21 | 38.77 | 17.95 | 84.1 |
| Brisbane | 7.099 | 2.528 | 6.682 | 3.435 | 13.18 |
| Cairo | 59.51 | 24.21 | 55.03 | 25.52 | 119 |
| Cantu | 37.46 | 15.68 | 34.47 | 15.81 | 76.18 |
| Charlotte | 15.83 | 9.452 | 13.58 | 4.59 | 39.99 |
| Chengdu | 73.64 | 30.27 | 68.21 | 31.45 | 148.3 |
| Cleveland | 22.62 | 13.35 | 19.49 | 6.771 | 56.89 |
| Coimbra | 28.82 | 17.47 | 24.55 | 8.444 | 74.61 |
| Colombo | 33.08 | 19.7 | 28.46 | 9.59 | 83.77 |
| Cordoba | 30.38 | 18.55 | 25.89 | 8.738 | 77.37 |
| Curitiba | 34.71 | 21.29 | 29.6 | 9.806 | 88.11 |
| Dallas | 22.37 | 13.33 | 19.28 | 6.608 | 56.69 |
| Detroit | 22.25 | 9.136 | 20.51 | 9.647 | 44.81 |
| Edmonton | 19.95 | 11.97 | 17.1 | 5.803 | 50.79 |
| Erfurt | 16.63 | 10.15 | 14.19 | 4.654 | 42.9 |
| Fairbanks | 19.79 | 8.408 | 18.22 | 8.27 | 40.29 |
| Florence | 46.14 | 27.48 | 39.54 | 13.63 | 117.7 |
| Fresno | 15.42 | 9.159 | 13.19 | 4.512 | 39.46 |
| Genoa | 35.03 | 20.98 | 30.1 | 10.07 | 89.42 |
| Glasgow | 19.29 | 12.04 | 16.42 | 5.37 | 50.16 |
| Halifax | 16.99 | 10.24 | 14.55 | 4.901 | 43.57 |
| Hangzhou | 73.79 | 44.4 | 63.14 | 21.76 | 185.2 |
| Hanoi | 52.03 | 21.63 | 48.08 | 22.25 | 105.5 |
| Helsinki | 14.1 | 4.318 | 13.46 | 7.46 | 24.32 |
| Hong Kong | 48.57 | 29.1 | 41.69 | 14.04 | 124.8 |
| Houston | 13 | 5.234 | 12.1 | 5.649 | 25.67 |
| Huelva | 28.72 | 17.69 | 24.58 | 8.194 | 73.49 |
| Hyderabad | 56.03 | 22.86 | 51.96 | 23.96 | 112.4 |
| Incheon | 54.09 | 32.17 | 46.45 | 15.52 | 138.6 |
| Indianapolis | 18.58 | 7.605 | 17.14 | 8.09 | 37.32 |
| Izmir | 56.7 | 34 | 48.66 | 16.51 | 145.5 |
| Jakarta | 48.4 | 29.02 | 41.72 | 13.88 | 121.7 |
| Jeddah | 62.4 | 38.19 | 53.48 | 18.26 | 159.8 |
| Karachi | 71.18 | 42.81 | 61.44 | 20.88 | 180.2 |
| Kuala Lumpur | 47.38 | 28.25 | 40.88 | 13.9 | 119.9 |
| Kuala Terengganu | 26.2 | 16.1 | 22.34 | 7.294 | 67.99 |
| Kuwait City | 48.82 | 29.95 | 41.44 | 13.82 | 128 |
| La Linea | 68.67 | 28.14 | 63.48 | 29.58 | 138.7 |
| Lahore | 26.66 | 15.97 | 22.84 | 7.789 | 68.49 |
| Las Vegas | 21.82 | 12.87 | 18.79 | 6.348 | 55.13 |
| Lijian | 62.58 | 36.99 | 54.26 | 18.18 | 157.1 |
| London | 22.4 | 13.78 | 19.15 | 6.212 | 57.74 |
| Los Angeles | 17.23 | 5.086 | 16.5 | 9.415 | 29.16 |
| Los Barrios | 24.71 | 14.93 | 21.16 | 7.17 | 62.56 |
| Lund | 11.89 | 7.236 | 10.17 | 3.439 | 30.85 |
| Manchester | 18.39 | 11.23 | 15.75 | 5.274 | 47.25 |
| Manila | 39.38 | 23.37 | 33.98 | 11.28 | 99.23 |
| Mantova | 31.99 | 13.31 | 29.6 | 13.5 | 65.07 |
| Marseille | 12.08 | 7.42 | 10.25 | 3.462 | 30.69 |
| Melbourne | 7.407 | 4.643 | 6.314 | 2.051 | 19.22 |
| Milan | 38.99 | 7.563 | 38.26 | 26.35 | 55.99 |
| Milwaukee | 23.8 | 14.3 | 20.39 | 6.838 | 61.56 |
| Montreal | 19.06 | 11.33 | 16.34 | 5.529 | 48.04 |
| Nagpur | 60.27 | 35.88 | 51.76 | 17.83 | 152.5 |
| Naples | 34.8 | 20.78 | 29.92 | 10.28 | 89.38 |
| New York | 17.69 | 7.223 | 16.38 | 7.591 | 35.57 |
| Orange, Texas | 11.19 | 6.723 | 9.614 | 3.201 | 28.77 |
| Oviedo | 32.08 | 19.14 | 27.67 | 9.314 | 81.41 |
| Pamplona | 29.35 | 12.17 | 27.16 | 12.57 | 59.19 |
| Paris | 12.98 | 5.51 | 11.94 | 5.383 | 26.4 |
| Pekanbaru | 43.77 | 25.85 | 37.78 | 12.77 | 109.8 |
| Phoenix | 22.82 | 13.6 | 19.63 | 6.743 | 57.4 |
| Pittsburg | 22.02 | 13.38 | 18.91 | 6.39 | 55.86 |
| Porto Alegre | 31.09 | 18.85 | 26.55 | 8.839 | 79.38 |
| Pullman | 19.85 | 11.91 | 17.03 | 5.8 | 50.37 |
| Rajshahi | 41.92 | 25.21 | 35.83 | 12.23 | 105 |
| Recife | 29.62 | 17.67 | 25.34 | 8.506 | 75 |
| Rio de Janeiro | 35.4 | 14.91 | 32.64 | 14.89 | 72.43 |
| Rochester | 14.29 | 8.895 | 12.21 | 4.044 | 37.21 |
| Sacramento | 17.43 | 7.11 | 16.14 | 7.489 | 34.63 |
| Salamanca | 16.23 | 9.895 | 13.89 | 4.723 | 41.89 |
| San Jose | 32.63 | 19.75 | 27.98 | 9.351 | 83.35 |
| San Joseusa | 18.12 | 7.424 | 16.75 | 7.72 | 36.41 |
| Santander | 34.66 | 14.13 | 32.13 | 14.75 | 69.61 |
| Santiago | 25.98 | 11.02 | 23.94 | 10.79 | 52.83 |
| Sao Paulo | 36.8 | 15.62 | 33.91 | 15.27 | 74.52 |
| Seattle | 16.55 | 5.559 | 15.7 | 8.246 | 29.83 |
| Seoul | 65.59 | 26.52 | 60.81 | 28.35 | 130.7 |
| Simi Valley | 14.92 | 6.051 | 13.87 | 6.391 | 29.67 |
| Skopje | 46.96 | 27.93 | 40.57 | 13.66 | 118.6 |
| St.Louis | 20.08 | 8.224 | 18.63 | 8.544 | 40.27 |
| Sydney | 7.159 | 3.114 | 6.551 | 2.947 | 14.83 |
| Taipei | 60.16 | 24.49 | 55.68 | 25.94 | 120.5 |
| Tartu | 50.66 | 30.2 | 43.4 | 14.38 | 127.6 |
| Thessaloniki | 45.1 | 26.99 | 38.71 | 12.93 | 114.9 |
| Tijuana | 15.93 | 9.487 | 13.68 | 4.658 | 40.69 |
| Tokyo | 27.88 | 16.77 | 23.86 | 8.119 | 70.71 |
| Toronto | 19.36 | 11.52 | 16.6 | 5.634 | 49.56 |
| Torontousa | 18.55 | 11.02 | 15.9 | 5.371 | 46.92 |
| Valenzuela | 50.81 | 30.23 | 43.83 | 14.84 | 125.3 |
| Wainuiomata | 4.439 | 2.782 | 3.771 | 1.192 | 11.75 |
| Washington DC | 15.53 | 4.014 | 15.01 | 9.112 | 24.86 |
| Windsor | 15.81 | 9.559 | 13.6 | 4.654 | 39.94 |
| Wrokalw | 53.48 | 31.66 | 46.16 | 15.8 | 132.4 |
| Zhengzhou | 80.12 | 48 | 68.71 | 23.03 | 204 |
| Zurich | 11.9 | 7.224 | 10.12 | 3.4 | 30.52 |

**Table E. Estimated expected posterior summaries of non-traffic-related PM_2.5_**

|  |  |  |  | 95% Bayesian interval | |
| --- | --- | --- | --- | --- | --- |
| City | Mean | St.Dev. | Median | 2.50% | 97.50% |
| Accra | 35.42 | 24.06 | 30.06 | 5.93 | 95.73 |
| Adelaide | 4.691 | 3.514 | 3.911 | 0.361 | 13.65 |
| Ahmedabad | 47.6 | 32.55 | 40.28 | 8.327 | 130.4 |
| Albacete | 22.16 | 15.47 | 18.56 | 4.126 | 61.57 |
| Alcala de Guadaira | 20.87 | 13.74 | 17.7 | 4.56 | 56.13 |
| Algeciras | 20.85 | 14.12 | 17.52 | 4.23 | 57.37 |
| Amsterdam | 10.36 | 4.583 | 9.502 | 3.978 | 21.73 |
| Antwerp | 10.1 | 6.646 | 8.537 | 2.245 | 27.14 |
| Arendtsville | 12.7 | 8.207 | 10.7 | 3.273 | 33.94 |
| Athens | 22.15 | 11.36 | 20.22 | 5.618 | 49.48 |
| Atlanta | 14.95 | 4.653 | 14.28 | 7.774 | 25.8 |
| Bakersfield | 12.61 | 8.14 | 10.67 | 3.182 | 33.52 |
| Baltimore | 12.34 | 8.267 | 10.44 | 2.459 | 33.34 |
| Bandung | 20.91 | 15.37 | 17.6 | 2.278 | 59.97 |
| Barcelona | 23.29 | 8.108 | 22.17 | 10.62 | 42.37 |
| Bari | 23.46 | 16.11 | 19.83 | 4.117 | 64.55 |
| Beijing | 55.22 | 14.12 | 53.74 | 32.23 | 87.42 |
| Belfast | 14.54 | 9.468 | 12.34 | 3.492 | 38.9 |
| Belo Horizonte | 24.13 | 18 | 20.37 | 1.044 | 69.08 |
| Birmingham | 11.96 | 5.476 | 10.9 | 4.364 | 25.5 |
| Birminghamusa | 15.19 | 6.634 | 13.96 | 6.051 | 31.26 |
| Brescia | 32.56 | 14.67 | 29.75 | 12.14 | 68.73 |
| Brisbane | 4.994 | 2.073 | 4.656 | 1.939 | 10.03 |
| Cairo | 43.15 | 20.1 | 39.57 | 14.49 | 92.74 |
| Cantu | 30.61 | 13.6 | 28.11 | 11.92 | 64.04 |
| Charlotte | 11.71 | 7.987 | 9.846 | 2.112 | 31.82 |
| Chengdu | 52.9 | 24.99 | 48.57 | 17.65 | 114.3 |
| Cleveland | 19.14 | 11.86 | 16.28 | 5.052 | 49.74 |
| Coimbra | 19.05 | 14.38 | 15.96 | 0.7579 | 55.83 |
| Colombo | 24.79 | 16.62 | 20.99 | 4.905 | 67.6 |
| Cordoba | 20 | 15.28 | 16.72 | 0.9416 | 58.05 |
| Curitiba | 22.15 | 17.35 | 18.53 | -0.314 | 65.93 |
| Dallas | 18.09 | 11.63 | 15.31 | 4.419 | 47.97 |
| Detroit | 18.8 | 8.129 | 17.19 | 7.695 | 38.51 |
| Edmonton | 16.73 | 10.58 | 14.22 | 4.338 | 43.91 |
| Erfurt | 13.68 | 8.91 | 11.57 | 3.291 | 36.57 |
| Fairbanks | 17.98 | 7.816 | 16.53 | 7.318 | 36.86 |
| Florence | 28.83 | 22.78 | 24.26 | -1.43 | 86.6 |
| Fresno | 12.77 | 8.062 | 10.85 | 3.264 | 33.71 |
| Genoa | 24.89 | 17.41 | 20.94 | 3.636 | 68.93 |
| Glasgow | 15.47 | 10.38 | 12.95 | 3.595 | 42.1 |
| Halifax | 14.34 | 9.09 | 12.16 | 3.677 | 37.89 |
| Hangzhou | 53.69 | 37.09 | 45.24 | 9.225 | 146.9 |
| Hanoi | 37.48 | 17.85 | 34.21 | 12.63 | 81.16 |
| Helsinki | 11.96 | 3.818 | 11.39 | 6.146 | 21.01 |
| Hong Kong | 33.68 | 24.04 | 28.31 | 3.837 | 95.09 |
| Houston | 10.44 | 4.498 | 9.629 | 4.161 | 21.51 |
| Huelva | 21.45 | 14.92 | 18 | 4.033 | 59.11 |
| Hyderabad | 41.36 | 19.02 | 37.99 | 14.62 | 88.49 |
| Incheon | 41.03 | 27.28 | 34.7 | 7.905 | 112.4 |
| Indianapolis | 14.92 | 6.562 | 13.65 | 5.861 | 31.03 |
| Izmir | 42.15 | 28.44 | 35.68 | 7.589 | 114.9 |
| Jakarta | 30.43 | 23.69 | 25.61 | -0.9712 | 88.72 |
| Jeddah | 46.2 | 32.24 | 38.86 | 8.632 | 126.3 |
| Karachi | 50.04 | 35.36 | 42.17 | 7.121 | 139.2 |
| Kuala Lumpur | 34.29 | 23.43 | 29.05 | 5.669 | 94.06 |
| Kuala Terengganu | 21.72 | 14.25 | 18.25 | 5.088 | 58.96 |
| Kuwait City | 41.94 | 26.97 | 35.26 | 10.8 | 112.7 |
| La Linea | 51.65 | 23.58 | 47.41 | 18.58 | 110.1 |
| Lahore | 20.49 | 13.61 | 17.27 | 4.3 | 55.88 |
| Las Vegas | 18.12 | 11.37 | 15.47 | 4.58 | 47.72 |
| Lijian | 48.35 | 31.58 | 41.13 | 10.68 | 129.4 |
| London | 15.99 | 11.48 | 13.41 | 2.184 | 45.09 |
| Los Angeles | 13.7 | 4.35 | 13.06 | 7.1 | 23.9 |
| Los Barrios | 19.74 | 12.9 | 16.66 | 4.61 | 52.72 |
| Lund | 9.962 | 6.407 | 8.42 | 2.543 | 26.54 |
| Manchester | 14.74 | 9.745 | 12.44 | 3.409 | 40.01 |
| Manila | 28.96 | 19.72 | 24.51 | 5.266 | 78.84 |
| Mantova | 25.83 | 11.48 | 23.73 | 9.776 | 54.58 |
| Marseille | 10.07 | 6.536 | 8.459 | 2.516 | 26.54 |
| Melbourne | 4.539 | 3.796 | 3.771 | -0.4287 | 14.01 |
| Milan | 27.83 | 6.265 | 27.24 | 17.27 | 41.79 |
| Milwaukee | 18.38 | 12.23 | 15.54 | 3.811 | 50.29 |
| Montreal | 15.51 | 9.95 | 13.1 | 3.783 | 40.99 |
| Nagpur | 45.12 | 30.34 | 38.07 | 8.953 | 122 |
| Naples | 19.18 | 16.98 | 16.29 | -6.209 | 60.47 |
| New York | 12.6 | 5.936 | 11.53 | 4.104 | 27.19 |
| Orange, Texas | 9.87 | 6.144 | 8.394 | 2.636 | 26.11 |
| Oviedo | 23.92 | 16.23 | 20.17 | 4.5 | 65.88 |
| Pamplona | 22.36 | 10.23 | 20.52 | 8.261 | 46.94 |
| Paris | 10.79 | 4.847 | 9.883 | 4.2 | 22.69 |
| Pekanbaru | 32.33 | 21.75 | 27.47 | 6.02 | 88.48 |
| Phoenix | 16.55 | 11.35 | 14.02 | 2.774 | 45.34 |
| Pittsburg | 17.28 | 11.47 | 14.6 | 3.923 | 46.34 |
| Porto Alegre | 20.49 | 15.39 | 17.21 | 0.9934 | 59.23 |
| Pullman | 17.73 | 10.97 | 15.07 | 4.891 | 45.98 |
| Rajshahi | 28.41 | 20.96 | 23.8 | 2.075 | 80 |
| Recife | 20.36 | 14.72 | 17.13 | 2.309 | 57.77 |
| Rio de Janeiro | 22.21 | 11.96 | 20.15 | 4.737 | 51.65 |
| Rochester | 12.31 | 8.036 | 10.45 | 3.133 | 32.97 |
| Sacramento | 14.21 | 6.168 | 13.05 | 5.662 | 29.27 |
| Salamanca | 13.68 | 8.803 | 11.57 | 3.509 | 36.55 |
| San Jose | 25.43 | 16.95 | 21.47 | 5.655 | 68.82 |
| San Joseusa | 15.03 | 6.511 | 13.78 | 5.987 | 31.17 |
| Santander | 26.76 | 11.97 | 24.52 | 10.06 | 56.39 |
| Santiago | 15.57 | 8.835 | 14.24 | 2.269 | 36.55 |
| Sao Paulo | 21.9 | 12.41 | 19.85 | 3.54 | 51.51 |
| Seattle | 12.98 | 4.742 | 12.24 | 5.956 | 24.29 |
| Seoul | 40.77 | 21.48 | 37.35 | 8.392 | 92.41 |
| Simi Valley | 12.91 | 5.442 | 11.92 | 5.324 | 26.33 |
| Skopje | 34.1 | 23.28 | 28.95 | 5.855 | 94.05 |
| St.Louis | 16.62 | 7.166 | 15.36 | 6.632 | 34.17 |
| Sydney | 4.662 | 2.517 | 4.2 | 1.121 | 10.75 |
| Taipei | 45.96 | 20.65 | 42.19 | 17.08 | 97.47 |
| Tartu | 40.49 | 26.16 | 34.25 | 9.392 | 107.9 |
| Thessaloniki | 27.7 | 21.76 | 23.35 | -2.114 | 81.84 |
| Tijuana | 12.73 | 8.201 | 10.79 | 3.01 | 33.95 |
| Tokyo | 20.84 | 14.21 | 17.46 | 3.791 | 56.75 |
| Toronto | 13.77 | 9.531 | 11.64 | 2.157 | 38.02 |
| Torontousa | 14.72 | 9.496 | 12.48 | 3.384 | 39 |
| Valenzuela | 39.45 | 26.07 | 33.57 | 8.815 | 104.4 |
| Wainuiomata | 3.62 | 2.445 | 3.043 | 0.8053 | 10 |
| Washington DC | 12.06 | 3.388 | 11.62 | 6.66 | 19.91 |
| Windsor | 13.47 | 8.549 | 11.48 | 3.57 | 34.72 |
| Wrokalw | 39.37 | 26.76 | 33.45 | 7.189 | 106 |
| Zhengzhou | 58.05 | 40.06 | 48.92 | 9.801 | 160.4 |
| Zurich | 10.17 | 6.454 | 8.57 | 2.624 | 27.1 |
